# Occupational risk of SARS-CoV-2 infection: a nationwide register-based study of the Danish workforce during the Covid-19 pandemic 2020-21

**DOI:** 10.1101/2022.10.25.22281247

**Authors:** Jens Peter Ellekilde Bonde, Luise Mølenberg Begtrup, Johan Høy Jensen, Esben Meulengracht Flachs, Vivi Schlünssen, Henrik Albert Kolstad, Kristina Jakobsson, Christel Nielsen, Kerstin Nilsson, Lars Rylander, Andreas Vilhelmsson, Kajsa Ugelvig Petersen, Sandra Søgaard Tøttenborg

## Abstract

**Objectives:** Most earlier studies on occupational risk of Covid-19 covering the entire workforce are based on relatively rare outcomes such as hospital admission and mortality. This study examines the incidence of SARS-CoV-2 infection by occupational group based upon real-time polymerase chain reaction tests (RT-PCR).

**Methods:** The cohort includes 2.4 million Danish employees, 20-69 years of age. All data were retrieved from public registries. The sex-specific incidence rate ratios (IRR) of first-occurring positive RT-PCR test from week 8 of 2020 through week 50 of 2021 were computed by Poisson regression for each 4-digit DISCO-08 job code with more than 100 employees (337 in men; 297 in women). Occupational groups with low risk of workplace infection according to a job exposure matrix constituted the reference group. Risk estimates were adjusted by demographic, social and health characteristics including household size, completed Covid-19 vaccination, pandemic wave and occupation-specific frequency of testing.

**Results:** IRR’s of SARS-CoV-2 infection were elevated in 34 occupations comprising 12 % of male employees and 45 occupations comprising 41 % of female employees. All IRR estimates were below 2.0. Decreased IRRs were observed in 85 occupations in men but none in women.

**Discussion:** We observed a modestly increased risk of SARS-CoV-2 infection among employees in numerous occupations indicating a large potential for preventive actions, especially in the female workforce. Cautious interpretation of observed risk in specific occupations is needed because of methodological issues inherent in analyses of RT-PCR-test results and because of multiple statistical tests.

**WHAT IS ALREADY KNOW ABOUT THIS TOPIC?:**

- Epidemiological studies suggest that the workplace contribute to the Covid-19 pandemic
- Results are mostly based upon studies of less frequent outcomes as Covid-19 morbidity or mortality which limits inference about risk in specific occupations

**WHAT THIS STUDY ADDS:**

- The risk of Covid-19 infection was increased in 34 of 337 occupations in men and in 45 of 297 occupations in women
- Some 12% of the Danish male workforce and 41% of the female workforce are at increased risk of Covid-19 infection

**HOW THIS RESEARCH MIGHT AFFECT RESEARCH, PRACTICE OR POLICY?:**

- Preventive actions targeting the workplace may contribute substantially to alleviate disease occurrence in the ongoing Covid-19 and similar future pandemics.

## INTRODUCTION

Less than three years have elapsed since the still ongoing Covid-19 pandemic took its start. During this short time span numerous epidemiological studies have consistently demonstrated that a range of occupations are associated with an increased risk of being infected with SARS-CoV-2 ^1-5^. The reasons may include higher frequencies of close social contacts at the workplace, caring for infected patients and less possibility to maintain sufficient distancing and efficiently use personal protective equipment ^6 7^. Other risk factors such as mode of transportation when commuting to work might also be of importance. Previously identified at-risk occupations include health care workers, teachers, bus and taxi drivers, meat packers, retail sale workers, bartenders, and police officers ^1-4 8^. The majority of studies have investigated risk across occupations using relatively rare outcomes such as Covid-19 related admission to hospital ^9-11^ or mortality ^4 9 10 12-16^. However, these approaches fail to address less severe non-hospital demanding cases and may thus underestimate risk of infection. Considering reports on delayed return to work ^17^ and of post Covid-19 conditions with persistent ill health following cases without acute critical disease ^18^, this is of importance not the least in occupations with many young and healthy employees who are less likely to be hospitalized but at equal or higher risk of infection.

We have recently reported increased occupational risk of severe Covid-19 in terms of Covid-19 related hospital admission among workers in the healthcare, social care, and transportation sectors ^8^. Based on the same cohort and time period, in this paper, we unravel the risk of asymptomatic and less severe Covid-19 using real-time polymerase chain reaction (RT-PCR) test as the outcome. Hereby, a strong gain is achieved in statistical power, which enables detection of potentially increased risk in occupations with too few (or too young and healthy) employees to allow analyses of hospital admissions. The capacity for free and on-demand Covid-19 RT-PCR testing covering all residents in Denmark was established during autumn 2020. By late spring 2021, the number of tests peaked at 0.8 million/week among 2.4 million employees (own data). While the cumulative incidence for Covid-19 related hospital admission in the workforce during the first almost two years of the pandemic in Denmark was 0.17% ^8^, the corresponding rate of at least one positive PCR test was 10.7% (own data).

The objective of this paper was to map the relative incidence rate of RT-PCR test positivity across all types of occupations during 2020-21 using a job-exposure-matrix derived reference group with an *a priori* assumed low level occupational risk of SARS-CoV-2 infection.

## METHODS

### Population and data

The study population was a nationwide register-based cohort of all Danish residents 20-69 years of age, who by December 31 of 2019 were gainfully employed (n = 2 451 542). It was extricated as a subset of the Danish Occupational Cohort with exposure data (DOC*X ^19^), which includes a range of demographic, social, occupational and health characteristics. Records were merged by the 10-digit unique personal identification number assigned by the Danish authorities. Additional information on vital data, SARS-CoV-2 tests, and Covid-19 vaccinations were provided by Statistics Denmark and the National Board of Health Data from baseline week 8, 2020 until end of follow-up week 50, 2021. Details on the cohort and its key variables have been published in a previous paper ^8^.

#### Occupational data

Jobs are classified by Statistics Denmark according to the Danish version of the International Standard Classification of Occupations (DISCO-08 ^20^), which is close to identical with the international classification ISCO-08 (423 4-digit codes, coverage 86% of all gainfully employed) and the Danish version of the Statistical Classification of Economic Activities in the European Communities (DB07) ^20^ (22 1.digit sections, 88 2-digit division, coverage > 99%).

#### Population characteristics

retrieved from Statistics Denmark by December 31 of 2019 included sex, age, duration of education in years, country of origin, hospital admission for one or more of eleven chronic diseases during 2010-2019, geographical residential area and the number of household members sharing the same residence including children. Information on health behaviors is not available in public registries with national coverage. We assigned sex-, age- and period-specific probability of current smoking and estimates of body mass index (kg/m^2^) by a Danish lifestyle JEM ^21^. These JEM derived variables predict mortality and ischemic heart disease independently of other risk factors in the Danish population of employees ^22^. Variables were grouped into the categories displayed in table 1.

**Table 1.**
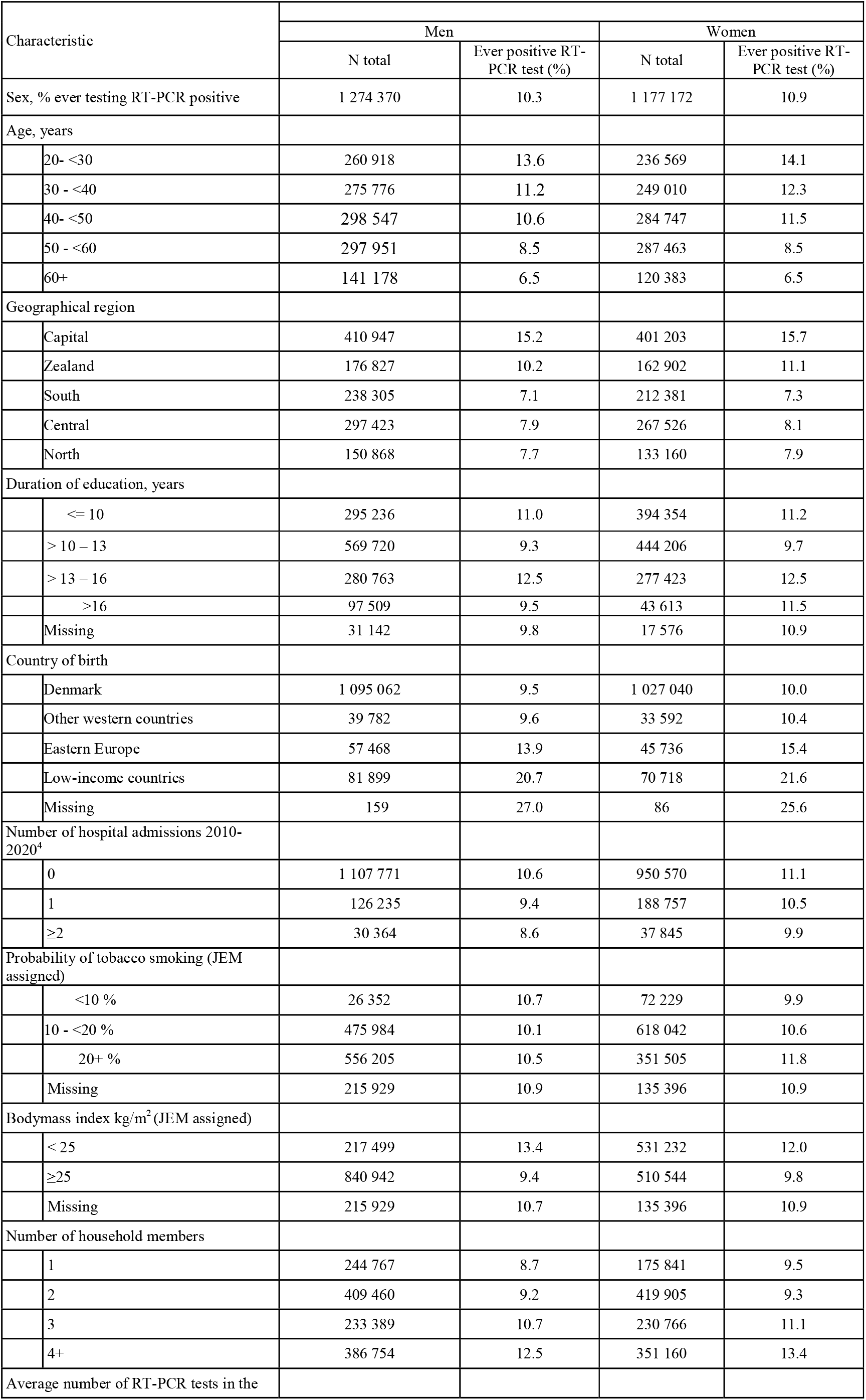
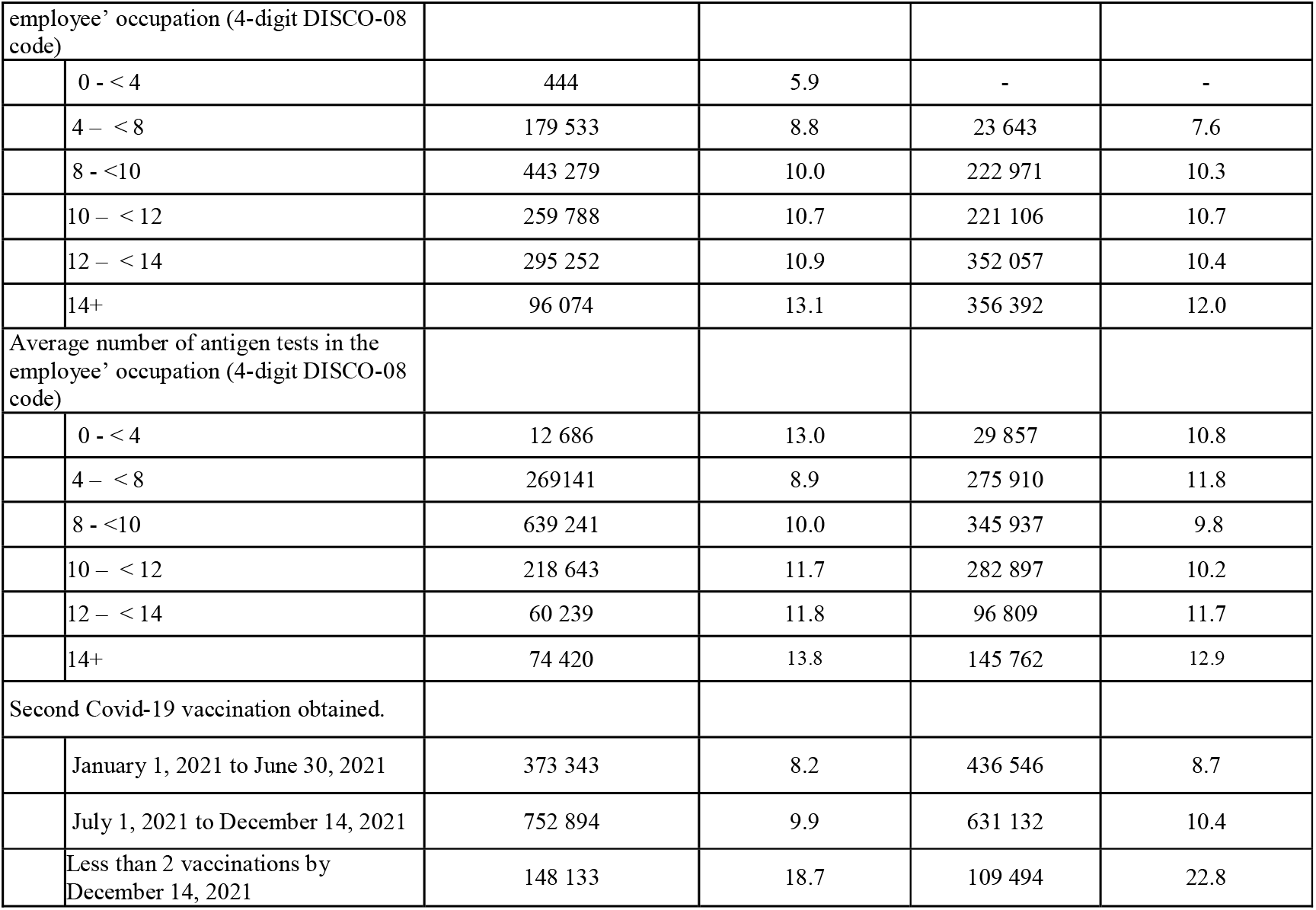
Prevalence of at least one positive SARS-CoV-2 RT-PCR test by characteristics of Danish male and female employees during the Covid-19 pandemic from week 8 of 2020 through week 52 of 2021.

#### SARS-CoV-2 test results

SARS-CoV-2 infection was identified by RT-PCR tests performed at Statens Serum Institut in Copenhagen or at an accredited hospital laboratory ^23^. The specificity of the RT-PCR test is extremely high (99.9%) ^24^, while the sensitivity in the general Danish population is unknown but believed to be above 80% ^24^. The sensitivity mainly depends on stage of infection and sampling procedure. In the early phase of the pandemic, RT-PCR tests were carried out for diagnostic purposes and for tracing of close contacts, but from autumn 2020 the test capacity was expanded and tests free of charge were offered to the entire population regardless of symptoms and contacts ^24^. Some 29.1 million tests were conducted in the study population until December 14^th^, 2021, of which 1.0 % were positive (own data).

#### Antigen tests (home tests)

have in particular been recommended in periods and geographical regions with a high load of viral transmission, and in age and occupational groups considered at high risk. The antigen test targets viral proteins and both sensitivity and specificity is much inferior compared to the RT-PCR test ^25^. Therefore, it is recommended that a positive antigen test is confirmed by a RT-PCR test. While antigen tests done as part of the nationwide and publicly funded test setup are included in the national database of test results, results of privately bought and used antigen tests were not. In this population only 25.5% persons with a positive antigen test after week 18 in 2021 (when the number of antigen tests peaked) had a RT-PCR test done within 2 days of which 72% were confirmed (own data).

#### Reference group

The reference group was defined *a priori* as occupations with employees working at home or not working with others according to an independent expert rated Covid-19 JEM ^6^. This reference group comprised 50 4-digit DISCO-08 job codes (n = 167 433 male and 201 969 female employees), mainly office clerks and administrative employees.

### Statistical analysis

We used Poisson regression to examine the sex-specific incidence rate ratios (IRR) of first-occurring positive SARS-CoV-2 RT-PCR test across all waves from week 8 of 2020 through week 50 of 2021 for each of 373 identified non-reference 4-digit DISCO-08 job codes with more than 100 employees (n=337 occupations for men and 297 for women). The time unit was one week, and weeks after the first positive RT-PCR test, death, emigration, or retirement were censored.

All analyses were run separately for men and women and risk estimates were adjusted by *a priori* selected characteristics regardless of association with exposures ^26^: age, duration of education, country of birth, geographical area, chronic disease, size of the household, body mass index and smoking. The covariate categories used in the statistical models were identical with those displayed in table 1. Completed Covid-19 vaccination (two doses with at least 21 days in between in the vast majority of cases) was entered into the statistical model as a time varying variable rather than censoring since vaccination does not entirely exclude the risk of infection. Finally, risk estimates were adjusted by the average RT-PCR and antigen test-frequency during the follow-up period for each 4-digit DISCO-08 occupational code (two separate variables) and pandemic wave defined as calendar period delimited by weeks with the lowest number of RT-PCR test between pandemic peaks (5 groups). Missing data for DISCO-08 (14%) and education (2%) were kept as separate categories in all analyses, which used a significance level of 0.05 and were carried out in SAS 9.4 (SAS Inst., Cary, NC, US) by access to a secured platform at Statistics Denmark.

## RESULTS

The study population included 2 451 542 employees who were traced for SARS-CoV-2 test results through 220 633 049 person weeks of follow-up. The RT-PCR testing frequency and the proportion of employees testing positive at least once varied strongly during the study period (figure 1), as did the average testing frequency across occupational groups (figure 2). The average number of RT-PCR tests per employee was 11.9 (median 8.0, range 0-223). In the entire study population, 10.7% tested positive at least once during the follow-up period (table 1). Among the 261 203 employees with a positive RT-PCR test, only 1917 (0.01%) had a second positive test later than 90 days (to exclude the possibility of ongoing first infection) after the first positive test (71% were tested on average 9.5 months later).

**Figure 1.**
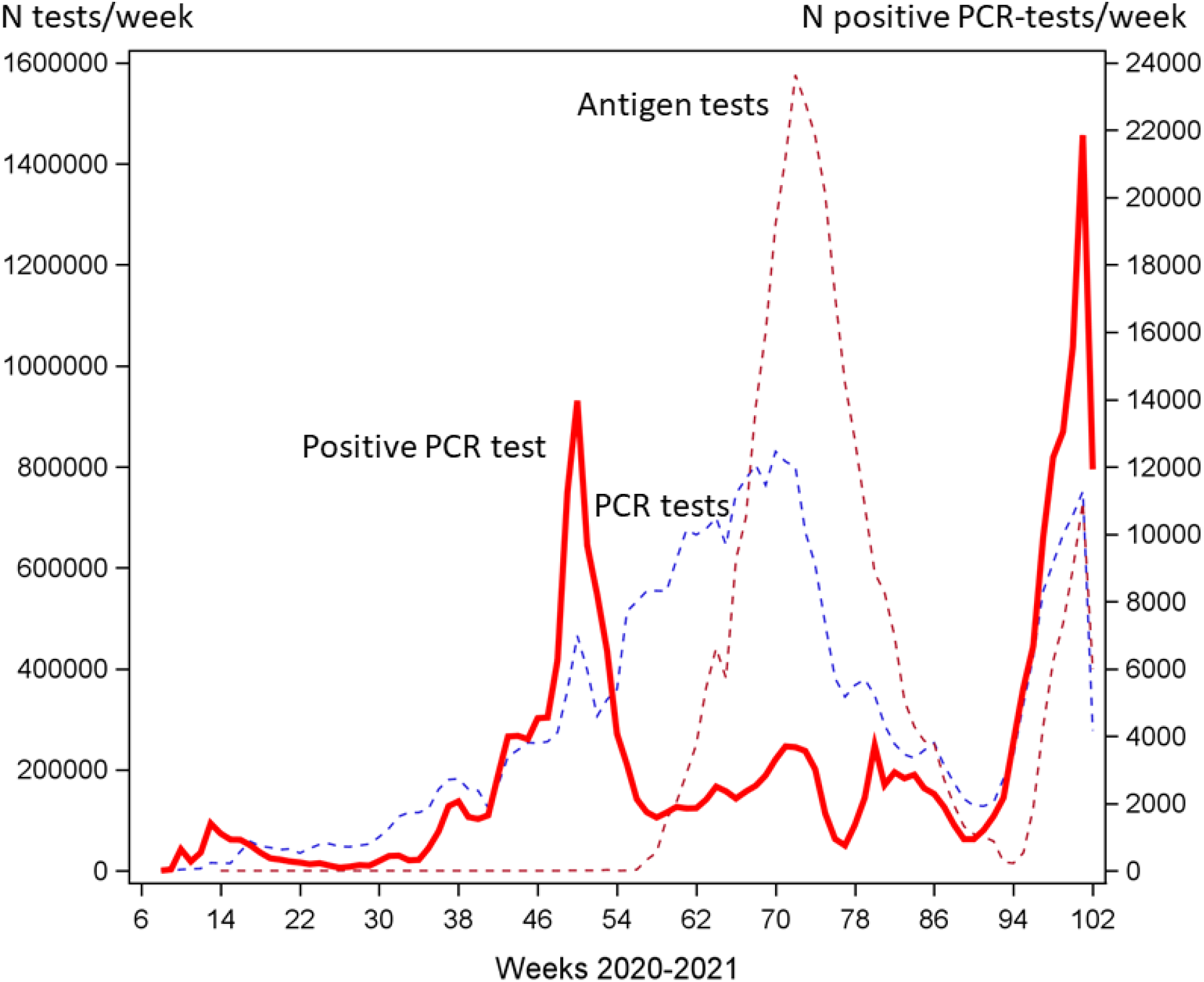
Weekly number of SARS-CoV-2 RT-PCR and antigen tests (left axis, dashed lines) and number of positive RT-PCR tests (right axis, solid line) among employees (n = 2 451 542) in Denmark from week 8 of 2020 through week 50 of 2021.

**Figure 2.**
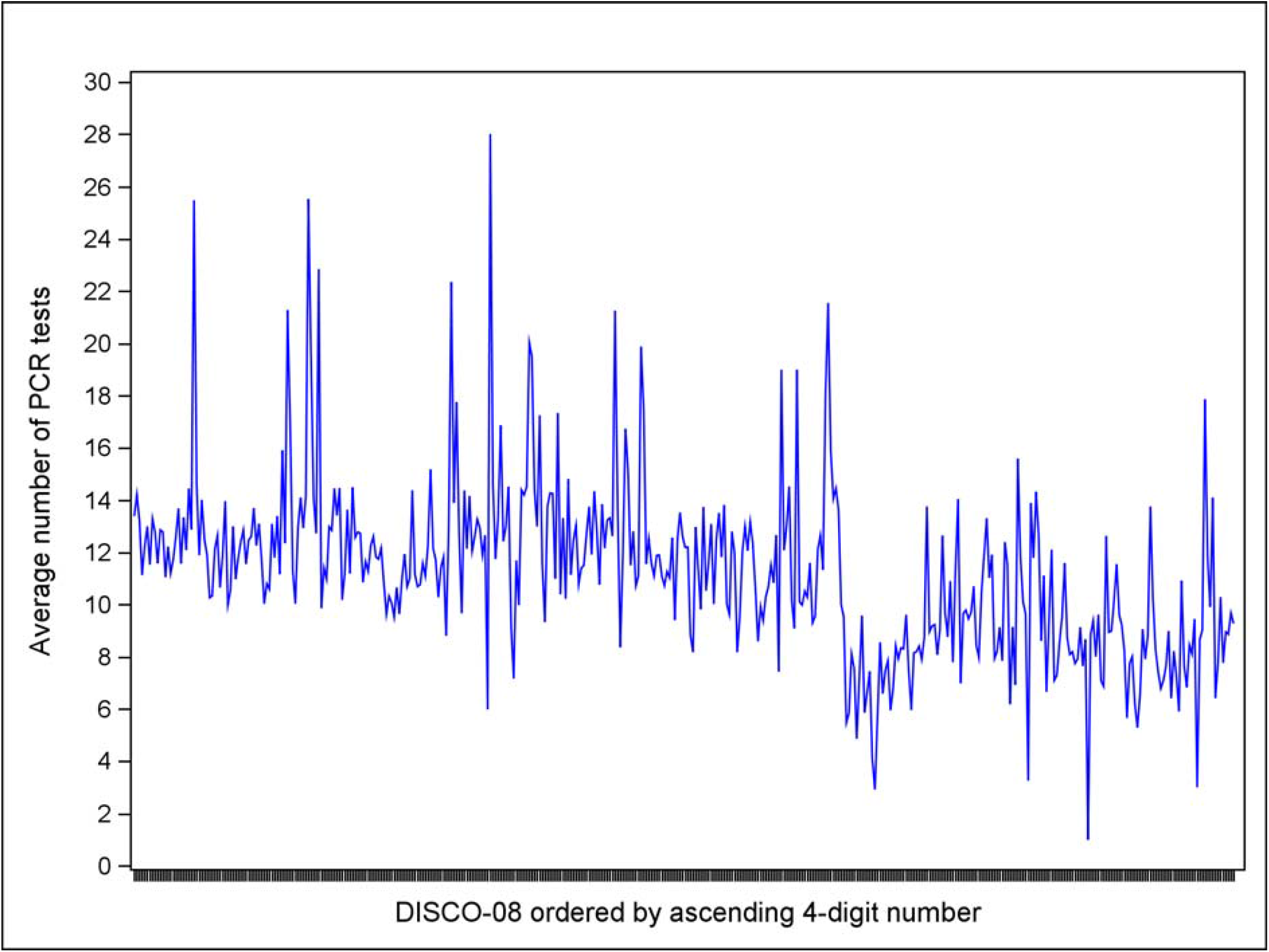
Distribution of average RT-PCR testing frequency across 423 4-digit DISCO-08 occupational groups during 2020-21.

While the infection rate was within the same range among men and women, it was strongly related to several covariates in both sexes – lower in the high age range and in employees with chronic disease, higher in the capital region, among foreign-born employees from non-western countries and among employees with high probability of low body mass index. Moreover, the infection rate increased with number of household members and with the average number of RT-PCR and antigen tests performed in the occupational group of the employee (table 1).

Among the 373 non-reference 4-digit DISCO-08 occupational codes with more than 100 employees (n = 1 742 280 employees with 156 909 762 weeks of follow-up), we observed an increased risk in 34 occupations in men and 45 in women. These occupations are listed in figure 3 and 4 and online supplemental table S1 and S2 grouped according to the 2-digit economic sector code (DB=07) with the highest number of employees within a given occupation. The corresponding figures for decreased risk were 86 occupational groups in men and none in women (online supplemental table S3). This indicates that 12% of the male workforce and 41% of the female workforce were at increased risk of SARS-CoV-2 infection given the selected reference group and the chosen statistical model. The magnitude of adjusted increased incidence rate ratios was ranging from 1.11 to 1.76 and of decreased risk from 0.42 to 0.90 (online supplemental tables 1-3).

**Figure 3.**
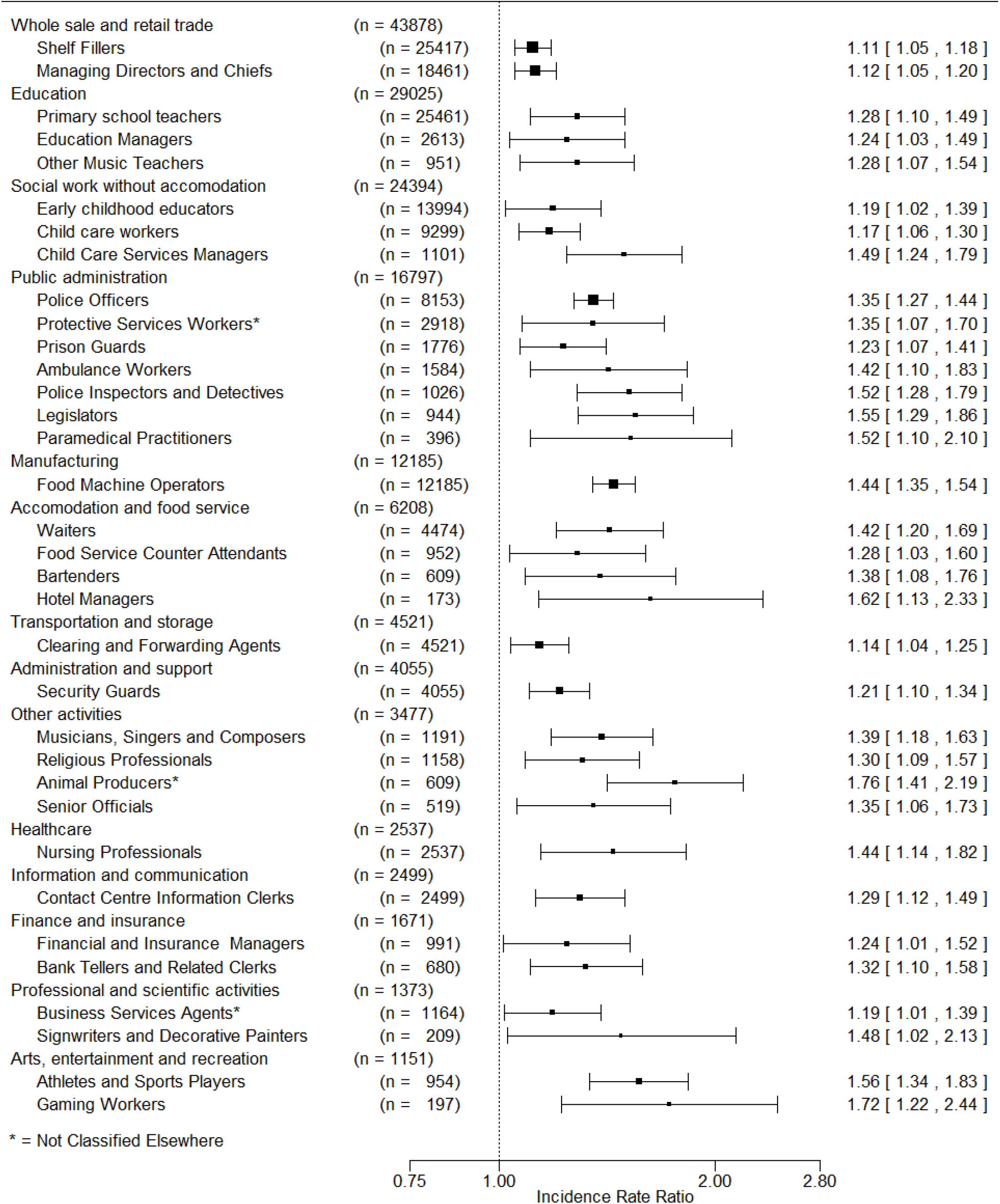
MEN. Adjusted^1^ incidence rate ratios above 1.0 and P-value < 0.05 for first positive SARS-CoV-2 RT-PCR test during the pandemic 2020-2021 by 4-digit DISCO-08 codes^2^. Ordered by main economic sector (2-digit DB07) and descending number of employees in occupational groups within economic sectors. ^1^ Adjusted for age, education, hospital admissions for chronic disease, country of birth, geographical region, number of household members, tobacco smoking, bodymass index, Covid19 vaccination, epidemic period and test frequency. ^2^ Reference is low-level exposed employees according a Covid-19 job exposure matrix.

**Figure 4.**
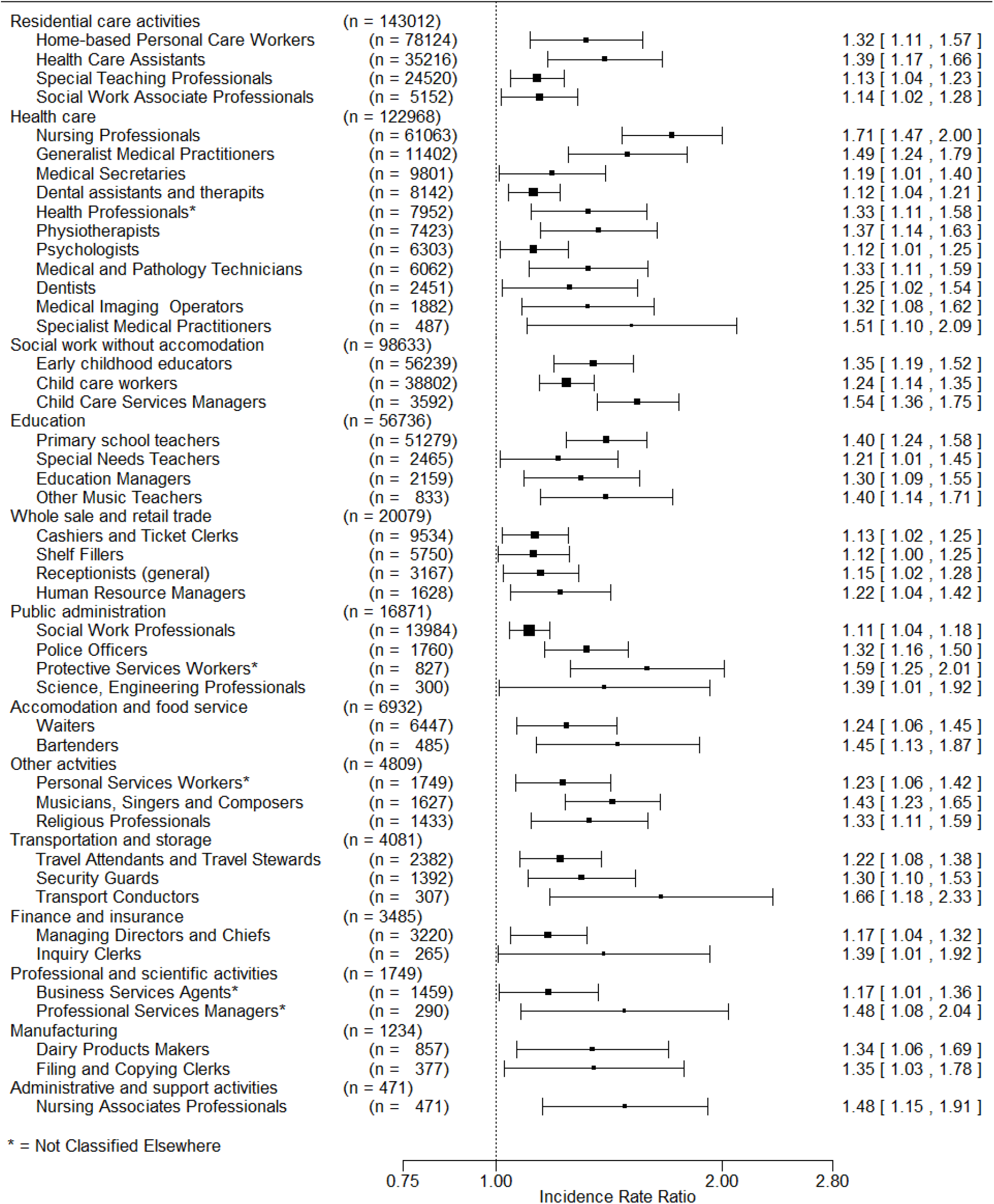
WOMEN. Adjusted^1^ incidence rate ratios (IRR) above 1.0 and P-value < 0.05 for first positive SARS-CoV-2 RT-PCR test during the pandemic 2020-2021 by 4-digit DISCO-08 codes^2^. Ordered by main economic sector (2-digit DB07) and descending number of employees in occupational groups within economic sectors. ^1^ Adjusted for age, education, hospital admissions for chronic disease, country of birth, geographical region, number of household members, tobacco smoking, bodymass index, Covid19 vaccination, epidemic period and test frequency. ^2^ Reference is low-level exposed employees according a Covid-19 job exposure matrix.

In men, the largest occupational groups with increased risk included shelf-fillers (retail store workers ensuring that shelves and product displays are stocked and often assisting as checkout operators) and primary school teachers. Other groups included day-care workers, police officers, prison guards, ambulance drivers, machine operators within food production, residential and healthcare workers, waiters and fast food operators (figure 3). Small groups, such as gaming workers (including bookmakers and casino croupiers), hotel managers, and animal producers were among those with highest relative risk. A range of health care occupations and bus- and taxi drivers did not appear with increased risk and no occupations within construction were represented.

Men in occupations with decreased risk were primarily employed within construction, manufacturing, and retail trade (online supplemental table S3)

In women, the largest occupations with increased risk were home-based personal care workers, healthcare assistants and social work associate professionals, and several specific groups within healthcare such as nurses, medical doctors and secretaries, laboratory technicians and imaging operators, dentists and dental assistants, physiotherapists, and psychologists. Also, teachers, receptionists, shelf fillers, and several other occupations were at increased risk (figure 4).

Adjustment for the occupation-specific testing frequency resulted in minor changes of risk estimates in either direction as did adjustment for the range of a priory selected covariates (online supplemental tables 1-3). In average, fully adjusted estimates were 1.1% higher than age-only adjusted estimates but with larger variation (SD 11.8 % of the mean difference).

## DISCUSSION

In this study of the entire Danish work force, the incidence rate ratio of SARS-CoV-2 infection during 2020-21 was elevated in 34 occupational groups comprising 12% of male employees and 45 occupations comprising 41% of female employees. All elevated adjusted IRR estimates were less than doubled. Decreased risk was observed in 86 occupations in men (31%) opposed to none in women. Increased risk was mainly apparent in health-, social- and childcare, education, and services entailing many and/or close contacts with people, but some occupations with increased risk were encountered in most major economic sectors.

The testing frequency was strongly skewed across occupational groups and the reasons for being tested likely also varied and may be related to both occupation, symptoms of Covid-19, and pandemic wave. Adjustment for the average occupation-specific test frequency at least partially accounts for increasing chance of a positive test with increasing number of tests performed but confounding due to varying indications for testing across occupations is not remedied. The result is unpredictable bias in either direction.

Nevertheless, among 368 697 employees in 33 4-digit DISCO-08 occupations that were found to be at increased risk of Covid-19 related hospital admission, 341 705 (93%) were also found to be at risk according to RT-PCR test results presented here. The largest occupations with increased adjusted risk of Covid-19 related hospital admission but not at increased adjusted risk of testing positive were bus and tram drivers and administrative and executive secretaries. Low testing frequencies in these occupations may contribute to this apparent inconsistency in spite of adjustment. For example, the average RT-PCR test frequency per employee was 7.9 among bus drivers compared to 11.9 across all occupations. Opposite, that numerous occupations (n = 49) were at increased risk of testing positive but not of Covid-19 related hospital admission is expected as the statistical power is much stronger for analysis of test results. These occupations include several specific jobs within healthcare and prison and security guards, police officers, waiters and bartenders, luggage porters, musicians, and many others. It should be acknowledged, however, that some of these associations may be random findings due to multiple testing and uncontrolled bias because of wide differences in reasons for testing and testing frequency. For example, the testing varied depending on the policy at different workplaces and for different professional groups. At the bottom line, unresolved limitations inherently related to analysis of testing results call for cautious interpretation of risk for specific occupations which must be construed in the light of findings in earlier studies applying other methodological approaches ^2 3 10 27-29^, and independent assessment of workplace risk factors - for instance using job-exposure matrices ^6 30^.

Restricting analyses to persons who ever performed a test during specified periods (labeled test-negative design ^31-33^) is not a promising option in this dataset with tremendous variation of test frequencies (figure 1 and 2). Conditioning on testing can result in collider bias and will inevitably produce spurious associations ^34^, which cannot be resolved unless all employees are tested repeatedly and systematically or tested completely at random ^35^.

Analyses were stratified by sex. While sex disparities with respect to determinants of severe Covid-19 are well established ^8 36^, there are no indications that one sex is substantial more susceptible to SARS-CoV-2 infection than the other which also is corroborated in this population (table 1). However, the extent of occupational exposure may differ between men and women within the same occupational group. Even at the 4-digit level, the DISCO-08 codes may include several different specific occupations. There are 423 4-digit codes but about 2200 specific occupational titles entailing different work tasks and potentially different risk of exposure. The sex distribution across these occupational titles within DISCO-08 groups may not be balanced.

Several occupational groups emerged with decreased risk of infection in men but not in women. It is conceivable that working may in some instances protect the employee from background infection if work is performed with limited contact to other people as for instance in lorry drivers (online supplemental table S3). However, the reduced risk within some occupations among men could also reflect that the JEM is less valid for men compared to women where no occupations were associated with significantly decreased risk. In a study of the same population, an alternative reference group for men was defined based upon outdoor work and with this reference few occupations were associated with decreased risk (unpublished work, personal communication).

Despite methodological limitations, the study corroborates most findings of occupationally increased risk of Covid-19 related hospital admissions and adds a number of occupational groups to the list of potentially at-risk jobs – at least partly due to higher statistical power allowing estimation of risk in more rare occupations and among younger and more healthy employees.

## CONCLUSION

The study corroborates some earlier findings on increased occupational risk of Covid-19 and indicates a modestly increased risk of SARS-CoV-2 infection among employees in several occupations that have not been identified in earlier studies using more rare outcomes. Methodological limitations call for cautious interpretation of risk for specific occupations which must be assessed in the light of findings in earlier studies. Nevertheless, the study indicates that risk of SARS-CoV-2 infection is far from is limited to healthcare, social care, and other essential occupations and that preventive action is warranted for a sizeable proportion of in particular the female workforce during possible forthcoming pandemics.

## Data Availability

The pseudonymized database used for the presented analyses is hosted by Statistics Denmark and is not publicly available. Permission to access the database can be granted by researchers at a research institution authorized by Statistics Denmark. On request the corresponding author can assist interested researchers to gain access.

## Acknowledgements

Læge Sofus Carl Emil Friis og Hustru Olga Doris Friis’ Legat and Interreg Øresund-Kattegat-Skagerrak (ÄrendeID: NYPS 20303383) are thanked for generous grants that proved crucial for undertaking this project.

## References

1. Murti M, Achonu C, Smith BT, et al. COVID-19 Workplace Outbreaks by Industry Sector and Their Associated Household Transmission, Ontario, Canada, January to June, 2020. Journal of occupational and environmental medicine 2021;63(7):574–80. doi: 10.1097/jom.0000000000002201 [published Online First: 2021/05/06]

2. van der Plaat DA, Madan I, Coggon D, et al. Risks of COVID-19 by occupation in NHS workers in England. Occup Environ Med 2022;79(3):176–83. doi: 10.1136/oemed-2021-107628 [published Online First: 2021/09/01]

3. Magnusson K, Nygård K, Methi F, et al. Occupational risk of COVID-19 in the first versus second epidemic wave in Norway, 2020. Euro Surveill 2021;26(40) doi: 10.2807/1560-7917.Es.2021.26.40.2001875 [published Online First: 2021/10/09]

4. Nafilyan V, Pawelek P, Ayoubkhani D, et al. Occupation and COVID-19 mortality in England: a national linked data study of 14.3 million adults. Occup Environ Med 2021 doi: 10.1136/oemed-2021-107818 [published Online First: 2021/12/31]

5. Rhodes S, Wilkinson J, Pearce N, et al. Occupational differences in SARS-CoV-2 infection: analysis of the UK ONS COVID-19 infection survey. J Epidemiol Community Health 2022;76(10):841–6. doi: 10.1136/jech-2022-219101 [published Online First: 20220711]

6. Oude Hengel KM, Burdorf A, Pronk A, et al. Exposure to a SARS-CoV-2 infection at work: development of an international job exposure matrix (COVID-19-JEM). Scand Journal Work, Environ Health 2022;48(1):61–70. doi: 10.5271/sjweh.3998 [published Online First: 2021/11/18]

7. Fadel M, Gilbert F, Legeay C, et al. Association between COVID-19 infection and work exposure assessed by the Mat-O-Covid job exposure matrix in the CONSTANCES cohort. Occup Environ Med 2022 doi: 10.1136/oemed-2022-108436 [published Online First: 20220920]

8. Bonde JPE, Sell L, Flachs EM, et al. Occupational risk of COVID-19 related hospital admission in Denmark 2020-2021: a follow-up study. Scandinavian journal of work, environment & health 2022 doi: 10.5271/sjweh.4063 [published Online First: 20221013]

9. Mutambudzi M, Niedwiedz C, Macdonald EB, et al. Occupation and risk of severe COVID-19: prospective cohort study of 120 075 UK Biobank participants. Occup Environ Med 2020 doi: 10.1136/oemed-2020-106731 [published Online First: 2020/12/11]

10. Chea N, Brown CJ, Eure T, et al. Risk Factors for SARS-CoV-2 Infection Among US Healthcare Personnel, May-December 2020. Emerg Infect Dis 2022;28(1):95–103. doi: 10.3201/eid2801.211803 [published Online First: 2021/12/03]

11. Alderling M, Albin M, Ahlbom A, Alfredsson L, Lystrøm J, Selander J. Risk att sjukhusvårdas för covid-19 i olika yrken. Rapport 2021:02, Stockholm: Centrum for arbets-och miljømedicin 2021

12. Chen YH, Glymour M, Riley A, et al. Excess mortality associated with the COVID-19 pandemic among Californians 18-65 years of age, by occupational sector and occupation: March through November 2020. PloS one 2021;16(6):e0252454. doi: 10.1371/journal.pone.0252454 [published Online First: 2021/06/05]

13. Windsor-Shellard BK J. Coronavirus (COVID-19) related deaths by occupation, England and Wales: deaths registered up to and including 20 April 2020. Statistical bulletin. London: Office for national statistics, 2020.

14. Hawkins D, Davis L, Kriebel D. COVID-19 deaths by occupation, Massachusetts, March 1-July 31, 2020. Am J Ind Med 2021 doi: 10.1002/ajim.23227 [published Online First: 2021/02/02]

15. Rimmer A. Covid-19: Two thirds of healthcare workers who have died were from ethnic minorities. BMJ (Clinical research ed) 2020;369:m1621. doi: 10.1136/bmj.m1621 [published Online First: 2020/04/25]

16. Billingsley S, Brandén M, Aradhya S, et al. COVID-19 mortality across occupations and secondary risks for elderly individuals in the household: A population register-based study. Scand J Work, Environ Health 2022;48(1):52–60. doi: 10.5271/sjweh.3992 [published Online First: 20211019]

17. Gualano MR, Rossi MF, Borrelli I, et al. Returning to work and the impact of post COVID-19 condition: A systematic review. Work 2022 doi: 10.3233/wor-220103 [published Online First: 20220801]

18. Franco JVA, Garegnani LI, Oltra GV, et al. Long-Term Health Symptoms and Sequelae Following SARS-CoV-2 Infection: An Evidence Map. Int J Environ Res Public Health 2022;19(16) doi: 10.3390/ijerph19169915 [published Online First: 20220811]

19. Flachs EM, Petersen SEB, Kolstad HA, et al. Cohort profile: DOC*X: a nationwide Danish occupational cohort with eXposure data – an open research resource. Int J Epidemiol 2019 doi: 10.1093/ije/dyz110 %J International Journal of Epidemiology

20. Statistik D. Dansk Branchekode DB07, v3: 2014-Copenhagen: Statistics Denmark; 2014 [Available from: https://www.dst.dk/da/Statistik/dokumentation/nomenklaturer/db07.

21. Bondo Petersen S, Flachs EM, Prescott EIB, et al. Job-exposure matrices addressing lifestyle to be applied in register-based occupational health studies. Occup Environ Med 2018;75(12):890–97. doi: 10.1136/oemed-2018-104991 [published Online First: 2018/09/03]

22. Bonde JPE, Flachs EM, Madsen IE, et al. Acute myocardial infarction in relation to physical activities at work: a nationwide follow-up study based on job-exposure matrices. Scand J Work, Environ Health 2020;46(3):268–77. doi: 10.5271/sjweh.3863 [published Online First: 2019/11/15]

23. Corman VM, Landt O, Kaiser M, et al. Detection of 2019 novel coronavirus (2019-nCoV) by real-time RT-PCR. Euro Surveill 2020;25(3) doi: 10.2807/1560-7917.Es.2020.25.3.2000045 [published Online First: 2020/01/30]

24. Vogels CBF, Brito AF, Wyllie AL, et al. Analytical sensitivity and efficiency comparisons of SARS-CoV-2 RT-qPCR primer-probe sets. Nat Microbiol 2020;5(10):1299–305. doi: 10.1038/s41564-020-0761-6 [published Online First: 2020/07/12]

25. Insitut SS. Falske antigen test Copenhagen 2021 [Available from: https://www.ssi.dk/aktuelt/nyheder/2021/antigentest-gav-47-falsk-negative-svar.

26. VanderWeele TJ. Principles of confounder selection. Eur J Epidemiol 2019;34(3):211–19. doi: 10.1007/s10654-019-00494-6 [published Online First: 2019/03/07]

27. Eyre DW, Lumley SF, O’Donnell D, et al. Differential occupational risks to healthcare workers from SARS-CoV-2 observed during a prospective observational study. Elife 2020;9 doi: 10.7554/eLife.60675 [published Online First: 2020/08/22]

28. Shah VP, Breeher LE, Alleckson JM, et al. Occupational exposure to severe acute respiratory coronavirus virus 2 (SARS-CoV-2) and risk of infection among healthcare personnel. Infect Control Hosp Epidemiol 2022:1–5. doi: 10.1017/ice.2021.533 [published Online First: 2022/01/07]

29. Dusefante A, Negro C, D’Agaro P, et al. Occupational Risk Factors for SARS-CoV-2 Infection in Hospital Health Care Workers: A Prospective Nested Case-Control Study. Life (Basel) 2022;12(2) doi: 10.3390/life12020263 [published Online First: 2022/02/26]

30. Baker MG, Peckham TK, Seixas NS. Estimating the burden of United States workers exposed to infection or disease: A key factor in containing risk of COVID-19 infection. PloS one 2020;15(4):e0232452. doi: 10.1371/journal.pone.0232452 [published Online First: 2020/04/29]

31. Vandenbroucke JP, Brickley EB, Pearce N, et al. The Evolving Usefulness of the Test-negative Design in Studying Risk Factors for COVID-19. Epidemiology (Cambridge, Mass) 2022;33(2):e7–e8. doi: 10.1097/ede.0000000000001438 [published Online First: 2021/11/21]

32. Vandenbroucke JP; Brickley EB; Vandenbroucke-Grauls CMJE; Pearce N. Test-negative design and matched case-control studies should be added to widespread testing of symptomatic persons for SARS-CoV-2, 2020.

33. Vandenbroucke JP, Brickley EB, Vandenbroucke-Grauls C, et al. A Test-Negative Design with Additional Population Controls Can Be Used to Rapidly Study Causes of the SARS-CoV-2 Epidemic. Epidemiology (Cambridge, Mass) 2020;31(6):836–43. doi: 10.1097/ede.0000000000001251 [published Online First: 2020/08/26]

34. Griffith GM T.T; Tudball M; Herbert A, et al. Collider bias undermines our understanding of Covid-19 disease risk and severity. Nat Commun 2020;11(1):5749. doi 10.1038/s41467-020-19478-2.

35. Burstyn I, Goldstein ND, Gustafson P. It can be dangerous to take epidemic curves of COVID-19 at face value. Can J Public Health 2020:1–4. doi: 10.17269/s41997-020-00367-6 [published Online First: 2020/06/25]

36. Yoshida Y, Chu S, Fox S, et al. Sex differences in determinants of COVID-19 severe outcomes - findings from the National COVID Cohort Collaborative (N3C). BMC Infect Dis 2022;22(1):784. doi: 10.1186/s12879-022-07776-7 [published Online First: 20221012]

