## Supplementary material for "Occupational risk of SARS-CoV-2 infection: a nationwide register-based study of the Danish workforce during the Covid-19 pandemic 2020-21": Suplemental tables S1-S3

**Running head:** Occupational risk of SARS-CoV-2 infection

**Authors: Authors:** Jens Peter Ellekilde Bonde^1,2^, Luise Mølenberg Begtrup^1,2^, Johan Høy Jensen^1^, Esben Meulengracht Flachs^1^, Vivi Schlünssen^3^, Henrik Albert Kolstad^4^, Kristina Jakobsson^5^, Christel Nielsen^6,7^, Kerstin Nilsson^6,8^, Lars Rylander^6^, Andreas Vilhelmsson^6^, Kajsa Ugelvig Petersen^1^, Sandra Søgaard Tøttenborg^1,2^

**ABSTRACT (WORD COUNT 246)**

**Objectives** Most earlier studies on occupational risk of Covid-19 covering the entire workforce are based on relatively rare outcomes such as hospital admission and mortality. This study examines the incidence of SARS-CoV-2 infection by occupational group based upon real-time polymerase chain reaction tests (RT-PCR).

**Methods** The cohort includes 2.4 million Danish employees, 20-69 years of age. All data were retrieved from public registries. The sex-specific incidence rate ratios (IRR) of first-occurring positive RT-PCR test from week 8 of 2020 through week 50 of 2021 were computed by Poisson regression for each 4-digit DISCO-08 job code with more than 100 employees (337 in men; 297 in women). Occupational groups with low risk of workplace infection according to a job exposure matrix constituted the reference group. Risk estimates were adjusted by demographic, social and health characteristics including household size, completed Covid-19 vaccination, pandemic wave and occupation-specific frequency of testing.

**Results** IRR’s of SARS-CoV-2 infection were elevated in 34 occupations comprising 12 % of male employees and 45 occupations comprising 41 % of female employees. All IRR estimates were below 2.0. Decreased IRRs were observed in 85 occupations in men but none in women.

**Discussion** We observed a modestly increased risk of SARS-CoV-2 infection among employees in numerous occupations indicating a large potential for preventive actions, especially in the female workforce. Cautious interpretation of observed risk in specific occupations is needed because of methodological issues inherent in analyses of RT-PCR-test results and because of multiple statistical tests.

Supplemental table S1. MEN AT INCREASED RISK in occupations with > 100 employees. Adjusted incidence rate ratios (IRR) above 1.0 and P-value < 0.05 for first positive SARS-CoV-2 RT-PCR test during the pandemic 2020-2021 in Denmark by 4-digit DISCO-08 codes^1^.

| **Main group economic code (DB07)** | | DISCO-08 code | Numbers | | % test positive | IRR^2^_age_ | IRR^3^  _age+ tests_ | IRR^4^_all_ | 95% CI | |
| --- | --- | --- | --- | --- | --- | --- | --- | --- | --- | --- |
|  | Occupation, 4-digit DISCO-08 codes  (descending number employees) |  | Em-ployees | RT-PCR tests |  |  |  |  |  |  |
| **Manufacturing,**  **N = 43 878** | | | | | | | | | | |
|  | Shelf Fillers | 9334 | 25417 | 215556 | 12.1 | 0.92 | 1.00 | 1.11 | 1.05 | 1.18 |
|  | Managing Directors and Chief Executives | 1120 | 18461 | 209652 | 9.5 | 1.17 | 1.25 | 1.12 | 1.05 | 1.20 |
| **Education,**  **N = 29 025** | | | | | | | | | | |
|  | Primary school teachers | 2341 | 25461 | 284665 | 13.3 | 1.37 | 1.13 | 1.28 | 1.10 | 1.49 |
|  | Education Managers | 1345 | 2613 | 29880 | 10.1 | 1.27 | 1.05 | 1.24 | 1.03 | 1.49 |
|  | Other Music Teachers | 2354 | 951 | 9016 | 13.5 | 1.28 | 1.23 | 1.28 | 1.07 | 1.54 |
| **Social Work Activities without Accommodation,**  **N = 24 394** | | | | | | | | | | |
|  | Early childhood educators | 2343 | 13994 | 164194 | 12.7 | 1.35 | 1.12 | 1.19 | 1.02 | 1.39 |
|  | Childcare workers | 5311 | 9299 | 94855 | 16.9 | 1.37 | 1.29 | 1.17 | 1.06 | 1.30 |
|  | Child Care Services Managers | 1341 | 1101 | 14193 | 10.4 | 1.48 | 1.38 | 1.49 | 1.24 | 1.79 |
| **Public Services, Defence and Security,**  **N = 16 797** | | | | | | | | | | |
|  | Police Officers | 5412 | 8153 | 114221 | 10.6 | 1.39 | 1.34 | 1.35 | 1.27 | 1.44 |
|  | Protective Services Workers Not Elsewhere Classified | 5419 | 2918 | 25754 | 15.6 | 1.34 | 1.59 | 1.35 | 1.07 | 1.70 |
|  | Prison Guards | 5413 | 1776 | 22597 | 10.4 | 1.29 | 1.39 | 1.23 | 1.07 | 1.41 |
|  | Ambulance Workers | 3258 | 1584 | 15721 | 13.5 | 1.33 | 1.58 | 1.42 | 1.10 | 1.83 |
|  | Police Inspectors and Detectives | 3355 | 1026 | 15405 | 9.4 | 1.62 | 1.74 | 1.52 | 1.28 | 1.79 |
|  | Legislators | 1111 | 944 | 9652 | 13.1 | 1.52 | 1.46 | 1.55 | 1.29 | 1.86 |
|  | Paramedical Practitioners | 2240 | 396 | 3929 | 16.8 | 1.67 | 1.98 | 1.52 | 1.10 | 2.10 |
| **Manufacturing,**  **N = 12 185** | | | | | | | | | | |
|  | Food and Related Products Machine Operators | 8160 | 12185 | 89144 | 22.8 | 1.64 | 1.76 | 1.44 | 1.35 | 1.54 |
| **Accommodation and Food Service Activities,**  **N = 6 208** | | | | | | | | | | |
|  | Waiters | 5131 | 4474 | 36198 | 22.3 | 1.42 | 1.18 | 1.42 | 1.20 | 1.69 |
|  | Food Service Counter Attendants | 5246 | 952 | 7145 | 23.0 | 1.23 | 1.03 | 1.28 | 1.03 | 1.60 |
|  | Bartenders | 5132 | 609 | 4732 | 23.7 | 1.41 | 1.17 | 1.38 | 1.08 | 1.76 |
|  | Hotel Managers | 1411 | 173 | 1925 | 15.6 | 1.76 | 1.69 | 1.62 | 1.13 | 2.33 |
| **Other Activities,**  **N = 21 284** | | | | | | | | | | |
|  | Clearing and Forwarding Agents | 3331 | 4521 | 61266 | 8.9 | 1.07 | 1.03 | 1.14 | 1.04 | 1.25 |
|  | Security Guards | 5414 | 4055 | 36671 | 14.5 | 1.23 | 1.32 | 1.21 | 1.10 | 1.34 |
|  | Nursing Professionals | 2221 | 2537 | 47498 | 8.0 | 1.58 | 1.88 | 1.44 | 1.14 | 1.82 |
|  | Contact Centre Information Clerks | 4222 | 2499 | 21190 | 16.3 | 1.03 | 0.97 | 1.29 | 1.12 | 1.49 |
|  | Musicians, Singers and Composers | 2652 | 1191 | 14769 | 11.1 | 1.46 | 1.41 | 1.39 | 1.18 | 1.63 |
|  | Business Services Agents Not Elsewhere Classified | 3339 | 1164 | 13314 | 12.3 | 1.25 | 1.20 | 1.19 | 1.01 | 1.39 |
|  | Religious Professionals | 2636 | 1158 | 12359 | 10.3 | 1.22 | 1.31 | 1.30 | 1.09 | 1.57 |
|  | Financial and Insurance Services Branch Managers | 1346 | 991 | 13859 | 8.3 | 1.21 | 1.30 | 1.24 | 1.01 | 1.52 |
|  | Athletes and Sports Players | 3421 | 954 | 19113 | 9.0 | 1.45 | 1.40 | 1.56 | 1.34 | 1.83 |
|  | Bank Tellers and Related Clerks | 4211 | 680 | 6782 | 18.1 | 1.45 | 1.39 | 1.32 | 1.10 | 1.58 |
|  | Animal Producers Not Elsewhere Classified | 6129 | 609 | 4103 | 20.7 | 1.26 | 1.35 | 1.76 | 1.41 | 2.19 |
|  | Senior Officials of Special Interest Organizations | 1114 | 519 | 6779 | 10.3 | 1.55 | 1.66 | 1.35 | 1.06 | 1.73 |
|  | Signwriters, Decorative Painters, Engravers and Etchers | 7316 | 209 | 1684 | 17.2 | 1.26 | 1.35 | 1.48 | 1.02 | 2.13 |
|  | Bookmakers, Croupiers and Related Gaming Workers | 4212 | 197 | 1667 | 19.8 | 1.43 | 1.37 | 1.72 | 1.22 | 2.44 |

^1^ Reference group: Employees with low likelihood of occupational SARS-CoV-2 exposure according to a Covid-19 job exposure matrix (sumscore for all eight rated indicators of SARS-CoV-2 workplace viral transmission = 0) ^6^

^2^ Adjusted for age (10 year groups)

^3^ In addition to age, adjusted for average number of RT-PCR tests and antigen tests in 4-digit-DISCO-08 codes (5 groups each)

^4^ In addition to age and test frequency, adjusted for duration of education at baseline (5 groups), number of hospital admissions for one or more of 11 chronic diseases in the 10 years preceding start of the pandemic (3 groups), country of birth (4 groups), geographical region (5 groups), number of household members (4 groups), probability of tobacco smoking (3 groups), bodymass index (2 groups), completed Covid19 vaccination (time varying variable, yes/no) and epidemic period (5 groups).

Supplemental table S2. WOMEN AT INCREASED RISK in occupations with > 100 employees. Adjusted incidence rate ratios (IRR) above 1.0 and P-value < 0.05 for first positive SARS-CoV-2 RT-PCR test during the pandemic 2020-2021 in Denmark by 4-digit DISCO-08 codes^1^.

| **Main group economic code (DB07)** | | DISCO-08 code | Numbers | | % test positive | IRR^2^_age_ | IRR^3^  _age+ tests_ | IRR^4^_all_ | 95% CI | |
| --- | --- | --- | --- | --- | --- | --- | --- | --- | --- | --- |
|  | Occupation, 4-digit DISCO-08 codes  (descending number employees) |  | Occupa-tion | RT-PCR tests |  |  |  |  |  |  |
| **Residential care activities,**  **N = 143 012** | | | | | | | | | | |
|  | Home-based Personal Care Workers | 5322 | 78124 | 1723537 | 13.1 | 1.46 | 1.72 | 1.32 | 1.11 | 1.57 |
|  | Health Care Assistants | 5321 | 35216 | 683509 | 12.6 | 1.39 | 1.64 | 1.39 | 1.17 | 1.66 |
|  | Special Teaching Professionals | 2357 | 24520 | 373326 | 9.9 | 1.09 | 1.13 | 1.13 | 1.04 | 1.23 |
|  | Social Work Associate Professionals | 3412 | 5152 | 57926 | 10.6 | 1.25 | 1.20 | 1.14 | 1.02 | 1.28 |
| **Healthcare,**  **N = 122 968** | | | | | | | | | | |
|  | Nursing Professionals | 2221 | 61063 | 1304617 | 12.4 | 1.39 | 1.62 | 1.71 | 1.47 | 2.00 |
|  | Generalist Medical Practitioners | 2211 | 11402 | 197310 | 11.9 | 1.23 | 1.44 | 1.49 | 1.24 | 1.79 |
|  | Medical Secretaries | 3344 | 9801 | 208144 | 9.0 | 1.02 | 1.19 | 1.19 | 1.01 | 1.40 |
|  | Dental assistants and therapits | 3251 | 8142 | 116684 | 12.3 | 1.25 | 1.30 | 1.12 | 1.04 | 1.21 |
|  | Health Professionals Not Elsewhere Classified | 2269 | 7952 | 185756 | 10.3 | 1.05 | 1.23 | 1.33 | 1.11 | 1.58 |
|  | Physiotherapists | 2264 | 7423 | 197788 | 11.4 | 1.17 | 1.37 | 1.37 | 1.14 | 1.63 |
|  | Psychologists | 2634 | 6303 | 98666 | 9.7 | 0.97 | 1.01 | 1.12 | 1.01 | 1.25 |
|  | Medical and Pathology Laboratory Technicians | 3212 | 6062 | 120460 | 11.4 | 1.27 | 1.48 | 1.33 | 1.11 | 1.59 |
|  | Dentists | 2261 | 2451 | 36949 | 10.5 | 1.14 | 1.32 | 1.25 | 1.02 | 1.54 |
|  | Medical Imaging and Equipment Operators | 3211 | 1882 | 40345 | 11.2 | 1.14 | 1.33 | 1.32 | 1.08 | 1.62 |
|  | Specialist Medical Practitioners | 2212 | 487 | 6726 | 10.9 | 1.49 | 1.73 | 1.51 | 1.10 | 2.09 |
| **Social Work Activities Without**  **Accommodation, N = 98 633** | | | | | | | | | | |
|  | Early childhood educators | 2343 | 56239 | 849991 | 13.4 | 1.42 | 1.32 | 1.35 | 1.19 | 1.52 |
|  | Childcare workers | 5311 | 38802 | 512204 | 13.5 | 1.42 | 1.37 | 1.24 | 1.14 | 1.35 |
|  | Child Care Services Managers | 1341 | 3592 | 53502 | 11.6 | 1.54 | 1.48 | 1.54 | 1.36 | 1.75 |
| **Education,**  **N = 56 736** | | | | | | | | | | |
|  | Primary school teachers | 2341 | 51279 | 710849 | 13.3 | 1.42 | 1.33 | 1.40 | 1.24 | 1.58 |
|  | Special Needs Teachers | 2352 | 2465 | 37801 | 11.4 | 1.32 | 1.24 | 1.21 | 1.01 | 1.45 |
|  | Education Managers | 1345 | 2159 | 26775 | 10.5 | 1.29 | 1.12 | 1.30 | 1.09 | 1.55 |
|  | Other Music Teachers | 2354 | 833 | 10894 | 12.2 | 1.33 | 1.24 | 1.40 | 1.14 | 1.71 |
| **Wholesale and retail trade,**  **N = 20 079** | | | | | | | | | | |
|  | Cashiers and Ticket Clerks | 5230 | 9534 | 104004 | 10.9 | 1.01 | 1.08 | 1.13 | 1.02 | 1.25 |
|  | Shelf Fillers | 9334 | 5750 | 62769 | 10.0 | 0.99 | 1.10 | 1.12 | 1.00 | 1.25 |
|  | Receptionists (general) | 4226 | 3167 | 42716 | 12.0 | 1.26 | 1.26 | 1.15 | 1.02 | 1.28 |
|  | Human Resource Managers | 1212 | 1628 | 21904 | 10.7 | 1.26 | 1.26 | 1.22 | 1.04 | 1.42 |
| **Public Service and Security**  **N =16 871** | | | | | | | | | | |
|  | Social Work and Counselling Professionals | 2635 | 13984 | 173292 | 10.1 | 1.03 | 1.03 | 1.11 | 1.04 | 1.18 |
|  | Police Officers | 5412 | 1760 | 29046 | 13.8 | 1.35 | 1.35 | 1.32 | 1.16 | 1.50 |
|  | Protective Services Workers Not Elsewhere Classified | 5419 | 827 | 9696 | 13.1 | 1.27 | 1.58 | 1.59 | 1.25 | 2.01 |
|  | Science and Engineering Professionals | 0210 | 300 | 4785 | 13.0 | 1.22 | 1.23 | 1.39 | 1.01 | 1.92 |
| **Accommodation and Food Service,**  **N = 6 932** | | | | | | | | | | |
|  | Waiters | 5131 | 6447 | 71505 | 15.4 | 1.30 | 1.32 | 1.24 | 1.06 | 1.45 |
|  | Bartenders | 5132 | 485 | 5454 | 18.8 | 1.62 | 1.63 | 1.45 | 1.13 | 1.87 |
| **Transportation and Storage,**  **N = 4 081** | | | | | | | | | | |
|  | Travel Attendants and Travel Stuarts | 5111 | 2382 | 32844 | 15.1 | 1.56 | 1.56 | 1.22 | 1.08 | 1.38 |
|  | Security Guards | 5414 | 1392 | 17320 | 12.7 | 1.34 | 1.49 | 1.30 | 1.10 | 1.53 |
|  | Transport Conductors | 5112 | 307 | 4008 | 14.3 | 1.82 | 2.11 | 1.66 | 1.18 | 2.33 |
| **Other activities,**  **N = 15 829** | | | | | | | | | | |
|  | Personal Services Workers Not Elsewhere Classified | 5169 | 1749 | 20134 | 13.9 | 1.42 | 1.32 | 1.23 | 1.06 | 1.42 |
|  | Musicians, Singers and Composers | 2652 | 1627 | 21692 | 12.0 | 1.38 | 1.39 | 1.43 | 1.23 | 1.65 |
|  | Religious Professionals | 2636 | 1433 | 17877 | 9.6 | 1.17 | 1.12 | 1.33 | 1.11 | 1.59 |
|  | Travel Attendants and Travel Stuarts | 5111 | 2382 | 32844 | 15.1 | 1.56 | 1.56 | 1.22 | 1.08 | 1.38 |
|  | Security Guards | 5414 | 1392 | 17320 | 12.7 | 1.34 | 1.49 | 1.30 | 1.10 | 1.53 |
|  | Transport Conductors | 5112 | 307 | 4008 | 14.3 | 1.82 | 2.11 | 1.66 | 1.18 | 2.33 |
|  | Managing Directors and Chief Executives | 1120 | 3220 | 39387 | 10.7 | 1.28 | 1.23 | 1.17 | 1.04 | 1.32 |
|  | Inquiry Clerks | 4225 | 265 | 3360 | 14.3 | 1.47 | 1.37 | 1.39 | 1.01 | 1.92 |
|  | Business Services Agents Not Elsewhere Classified | 3339 | 1459 | 18525 | 11.9 | 1.25 | 1.25 | 1.17 | 1.01 | 1.36 |
|  | Professional Services Managers Not Elsewhere Classified | 1349 | 290 | 3903 | 13.4 | 1.55 | 1.61 | 1.48 | 1.08 | 2.04 |
|  | Dairy Products Makers | 7513 | 857 | 10038 | 9.2 | 0.95 | 1.06 | 1.34 | 1.06 | 1.69 |
|  | Filing and Copying Clerks | 4415 | 377 | 4894 | 16.7 | 1.47 | 1.39 | 1.35 | 1.03 | 1.78 |
|  | Nursing Associates Professionals | 3221 | 471 | 8886 | 20.0 | 1.69 | 1.97 | 1.48 | 1.15 | 1.91 |

^1^ Reference group: Employees with low likelihood of occupational SARS-CoV-2 exposure according to a Covid-19 job exposure matrix (sumscore for all eight rated indicators of SARS-CoV-2 workplace viral transmission = 0) ^6^

^2^ Adjusted for age (10 year groups)

^3^ In addition to age, adjusted for average number of RT-PCR tests and antigen tests in 4-digit-DISCO-08 codes (5 groups each)

^4^ In addition to age and test frequency, adjusted for duration of education at baseline (5 groups), number of hospital admissions for one or more of 11 chronic diseases in the 10 years preceding start of the pandemic (3 groups), country of birth (4 groups), geographical region (5 groups), number of household members (4 groups), probability of tobacco smoking (3 groups), bodymass index (2 groups), completed Covid19 vaccination (time varying variable, yes/no) and epidemic period (5 groups).

Supplemental table S3. MEN AT DECREASED RISK in occupations with > 100 employees. Adjusted incidence rate ratios (IRR) < 1.0 and p-value < 0.05 for first positive SARS-CoV-2 RT-PCR test during the pandemic 2020-2021 in Denmark by 4-digit DISCO-08 codes^1^.

| **Main group economic code (DB07)** | | DISCO-08 code | Numbers | | % test positive | IRR^2^_age_ | IRR^3^_age+ tests_ | IRR^4^_all_ | 95% CI | |
| --- | --- | --- | --- | --- | --- | --- | --- | --- | --- | --- |
| Occupation, 4-digit DISCO-08 codes  (descending number employees) | |  | Occupa-tion | RT-PCR tests |  |  |  |  |  |  |
| **Construction,**  **N = 113 421** | | | | | | | | | | |
|  | Carpenters and Joiners | 7115 | 29605 | 218163 | 9.0 | 0.76 | 0.82 | 0.74 | 0.70 | 0.78 |
|  | Building and Related Electricians | 7411 | 23141 | 239300 | 9.5 | 0.83 | 0.90 | 0.78 | 0.72 | 0.84 |
|  | Civil Engineering Labourers | 9312 | 21485 | 154814 | 6.3 | 0.62 | 0.74 | 0.54 | 0.43 | 0.67 |
|  | Building Construction Labourers | 9313 | 10880 | 71774 | 8.3 | 0.75 | 0.89 | 0.58 | 0.46 | 0.72 |
|  | Bricklayers and Related Workers | 7112 | 10166 | 56379 | 8.0 | 0.70 | 0.83 | 0.54 | 0.43 | 0.68 |
|  | Painters and Related Workers | 7131 | 6352 | 44060 | 10.1 | 0.91 | 0.98 | 0.73 | 0.66 | 0.80 |
|  | Earthmoving and Related Plant Operators | 8342 | 3313 | 20837 | 5.0 | 0.48 | 0.56 | 0.45 | 0.35 | 0.59 |
|  | Building Frame and Related Trades Workers Not Elsewhere Classified | 7119 | 3217 | 24555 | 9.1 | 0.86 | 1.02 | 0.64 | 0.50 | 0.82 |
|  | Concrete Placers, Concrete Finishers and Related Workers | 7114 | 1366 | 8795 | 7.3 | 0.64 | 0.69 | 0.59 | 0.48 | 0.73 |
|  | Floor Layers and Tile Setters | 7122 | 1346 | 8875 | 10.0 | 0.87 | 0.94 | 0.76 | 0.64 | 0.91 |
|  | Roofers | 7121 | 1193 | 6956 | 7.5 | 0.65 | 0.70 | 0.59 | 0.47 | 0.73 |
|  | Insulation Workers | 7124 | 892 | 6966 | 7.8 | 0.73 | 0.87 | 0.58 | 0.42 | 0.80 |
|  | Well Drillers and Borers and Related Workers | 8113 | 465 | 5648 | 6.5 | 0.63 | 0.75 | 0.62 | 0.41 | 0.93 |
| **Manufacturing,**  **N=86 326** | | | | | | | | | | |
|  | Engineering Professionals Not Elsewhere Classified | 2149 | 7369 | 87879 | 8.0 | 0.76 | 0.82 | 0.79 | 0.72 | 0.86 |
|  | Industrial and Production Engineers | 2141 | 6756 | 82368 | 7.2 | 0.67 | 0.73 | 0.72 | 0.66 | 0.79 |
|  | Mechanical Engineering Technicians | 3115 | 6358 | 79336 | 8.2 | 0.81 | 0.87 | 0.84 | 0.76 | 0.92 |
|  | Physical and Engineering Science Technicians | 3119 | 5785 | 70301 | 7.7 | 0.74 | 0.80 | 0.81 | 0.73 | 0.89 |
|  | Mechanical Engineers | 2144 | 5265 | 64776 | 8.1 | 0.78 | 0.84 | 0.80 | 0.73 | 0.89 |
|  | Metal Processing Plant Operators | 8121 | 4745 | 32307 | 6.5 | 0.63 | 0.75 | 0.58 | 0.45 | 0.73 |
|  | Management and Organization Analysts | 2421 | 4665 | 52626 | 10.5 | 0.98 | 0.94 | 0.89 | 0.81 | 0.98 |
|  | Toolmakers and Related Workers | 7222 | 4341 | 39324 | 6.6 | 0.60 | 0.71 | 0.75 | 0.59 | 0.95 |
|  | Plastic Products Machine Operators | 8142 | 4095 | 30838 | 8.8 | 0.86 | 1.02 | 0.74 | 0.59 | 0.95 |
|  | Electronics Engineers | 2152 | 3447 | 40060 | 8.3 | 0.78 | 0.93 | 0.71 | 0.55 | 0.92 |
|  | Wood Processing Plant Operators | 8172 | 3149 | 20398 | 5.9 | 0.58 | 0.69 | 0.58 | 0.45 | 0.75 |
|  | Draughts persons | 3118 | 3063 | 34330 | 7.7 | 0.73 | 0.78 | 0.80 | 0.70 | 0.92 |
|  | Sheet Metal Workers | 7213 | 3038 | 23657 | 7.9 | 0.73 | 0.86 | 0.64 | 0.50 | 0.82 |
|  | Electrical Mechanics and Fitters | 7412 | 2924 | 34493 | 7.0 | 0.71 | 0.85 | 0.65 | 0.51 | 0.84 |
|  | Chemical Engineers | 2145 | 2853 | 33137 | 10.0 | 0.96 | 1.04 | 0.83 | 0.72 | 0.95 |
|  | Chemical and Physical Science Technicians | 3111 | 2525 | 30067 | 9.0 | 0.86 | 0.92 | 0.77 | 0.67 | 0.88 |
|  | Mechanical Machinery Assemblers | 8211 | 2268 | 21875 | 7.5 | 0.75 | 0.89 | 0.70 | 0.54 | 0.92 |
|  | Bakers, Pastry-cooks and Confectionery Makers | 7512 | 2212 | 14889 | 7.4 | 0.67 | 0.72 | 0.81 | 0.69 | 0.95 |
|  | Electrical and Electronic Equipment Assemblers | 8212 | 1949 | 20337 | 8.9 | 0.87 | 1.04 | 0.74 | 0.57 | 0.96 |
|  | Pharmacists | 2262 | 1538 | 17384 | 10.7 | 1.00 | 1.08 | 0.83 | 0.69 | 0.99 |
|  | Woodworking Machine Tool Setters and Operators | 7523 | 1392 | 9104 | 4.8 | 0.47 | 0.55 | 0.49 | 0.35 | 0.67 |
|  | Cement, Stone and Other Mineral Products Machine Operators | 8114 | 1348 | 9108 | 7.8 | 0.76 | 0.90 | 0.69 | 0.51 | 0.92 |
|  | Mathematicians, Actuaries, and Statisticians | 2120 | 1275 | 12683 | 9.6 | 0.90 | 0.86 | 0.78 | 0.65 | 0.94 |
|  | Craft and Related Workers Not Elsewhere Classified | 7549 | 1011 | 10974 | 7.2 | 0.73 | 0.79 | 0.70 | 0.55 | 0.90 |
|  | Printers | 7322 | 784 | 6474 | 5.6 | 0.55 | 0.65 | 0.67 | 0.47 | 0.97 |
|  | Chemists | 2113 | 641 | 8199 | 8.1 | 0.76 | 0.82 | 0.67 | 0.51 | 0.89 |
|  | Print Finishing and Binding Workers | 7323 | 590 | 4599 | 7.5 | 0.76 | 0.89 | 0.67 | 0.46 | 0.96 |
|  | Metal Polishers, Wheel Grinders and Tool Sharpeners | 7224 | 528 | 3994 | 8.0 | 0.78 | 0.93 | 0.64 | 0.44 | 0.93 |
|  | Glass and Ceramics Plant Operators | 8181 | 412 | 2592 | 6.1 | 0.59 | 0.70 | 0.51 | 0.33 | 0.80 |
| **Wholesale, Retail and Vehicle Repair,**  **N = 60 921** | | | | | | | | | | |
|  | Shop Sales Assistants | 5223 | 27908 | 244374 | 12.2 | 0.95 | 0.91 | 0.85 | 0.81 | 0.91 |
|  | Motor Vehicle Mechanics and Repairers | 7231 | 17752 | 158107 | 8.1 | 0.71 | 0.77 | 0.90 | 0.84 | 0.96 |
|  | Retail and Wholesale Trade Managers | 1420 | 4597 | 45042 | 10.1 | 0.90 | 0.97 | 0.83 | 0.74 | 0.93 |
|  | Electronics Engineering Technicians | 3114 | 4327 | 52166 | 7.6 | 0.75 | 0.89 | 0.65 | 0.52 | 0.83 |
|  | Butchers, Fishmongers and Related Food Preparers | 7511 | 2379 | 17011 | 8.9 | 0.74 | 0.80 | 0.70 | 0.61 | 0.82 |
|  | Spray Painters and Varnishers | 7132 | 2004 | 11392 | 6.8 | 0.60 | 0.72 | 0.50 | 0.38 | 0.65 |
|  | Vehicle Cleaners | 9122 | 1081 | 7755 | 11.0 | 0.99 | 1.17 | 0.72 | 0.54 | 0.95 |
|  | Information and Communications Technology Installers and Servicers | 7422 | 489 | 5734 | 7.0 | 0.63 | 0.68 | 0.64 | 0.46 | 0.91 |
|  | Precision-instrument Makers and Repairers | 7311 | 384 | 3982 | 9.4 | 0.86 | 0.93 | 0.67 | 0.48 | 0.95 |
| **Information and Communication,**  **N =41 182** | | | | | | | | | | |
|  | Software Developers | 2512 | 19786 | 198924 | 8.9 | 0.82 | 0.88 | 0.77 | 0.72 | 0.83 |
|  | Information and Communications Technology User Support | 3512 | 6216 | 64160 | 9.1 | 0.79 | 0.86 | 0.75 | 0.68 | 0.83 |
|  | Systems Administrators | 2522 | 3381 | 36126 | 8.7 | 0.84 | 0.90 | 0.81 | 0.72 | 0.92 |
|  | Software and Applications Developers and Analysts Not Elsewhere | 2519 | 3169 | 33005 | 9.2 | 0.85 | 0.92 | 0.79 | 0.70 | 0.90 |
|  | Web and Multimedia Developers | 2513 | 2269 | 21561 | 9.3 | 0.78 | 0.75 | 0.76 | 0.66 | 0.88 |
|  | Database and Network Professionals Not Elsewhere Classified | 2529 | 2249 | 22867 | 9.7 | 0.91 | 0.98 | 0.82 | 0.71 | 0.94 |
|  | Computer Network and Systems Technicians | 3513 | 1819 | 20241 | 8.0 | 0.76 | 0.82 | 0.72 | 0.61 | 0.85 |
|  | Information and Communications Technology Operations | 3511 | 1284 | 14018 | 8.5 | 0.82 | 0.88 | 0.73 | 0.60 | 0.89 |
|  | Computer Network Professionals | 2523 | 820 | 9811 | 9.8 | 0.88 | 0.95 | 0.80 | 0.64 | 1.00 |
|  | Web Technicians | 3514 | 189 | 1902 | 6.3 | 0.53 | 0.51 | 0.51 | 0.29 | 0.89 |
| **Administrative and Support Activities,**  **N = 35 497** | | | | | | | | | | |
|  | Building caretakers | 5153 | 16946 | 211688 | 8.5 | 0.94 | 1.01 | 0.84 | 0.79 | 0.91 |
|  | Cleaners and Helpers in Offices, Hotels and Other Establishments | 9112 | 13568 | 105501 | 13.9 | 1.31 | 1.56 | 0.78 | 0.63 | 0.98 |
|  | Gardeners Horticultural and Nursery Growers | 6113 | 3710 | 26930 | 6.9 | 0.62 | 0.74 | 0.68 | 0.53 | 0.86 |
|  | Other Cleaning Workers | 9129 | 1273 | 7622 | 11.1 | 0.99 | 1.18 | 0.63 | 0.48 | 0.83 |
| **Transportation and storage,**  **N = 35 058** | | | | | | | | | | |
|  | Heavy Truck and Lorry Drivers | 8332 | 22745 | 136300 | 5.9 | 0.57 | 0.68 | 0.52 | 0.42 | 0.66 |
|  | Car, Taxi and Van Drivers | 8322 | 5835 | 43741 | 12.2 | 1.20 | 1.41 | 0.78 | 0.62 | 0.99 |
|  | Messengers, Package Deliverers and Luggage Porters | 9621 | 2748 | 18934 | 13.9 | 1.26 | 1.36 | 0.89 | 0.79 | 0.99 |
|  | Ships’ Engineers | 3151 | 1803 | 21032 | 6.1 | 0.58 | 0.62 | 0.68 | 0.56 | 0.83 |
|  | Garbage and Recycling Collectors | 9611 | 1182 | 7309 | 11.7 | 1.11 | 1.31 | 0.74 | 0.57 | 0.98 |
| **Other Activities,**  **N = 20 116** | | | | | | | | | | |
|  | Armed Forces Occupations, Other Ranks | 0310 | 7930 | 101903 | 10.4 | 0.79 | 0.75 | 0.84 | 0.74 | 0.96 |
|  | Philosophers, Historians and Political Scientists | 2633 | 1053 | 10566 | 8.7 | 0.82 | 0.88 | 0.67 | 0.54 | 0.84 |
|  | Physicists and Astronomers | 2111 | 509 | 5215 | 7.7 | 0.70 | 0.68 | 0.72 | 0.52 | 0.99 |
|  | Sociologists, Anthropologists and Related Professionals | 2632 | 428 | 4311 | 7.5 | 0.65 | 0.61 | 0.66 | 0.46 | 0.94 |
|  | Field Crop and Vegetable Growers | 6111 | 1455 | 7271 | 7.6 | 0.64 | 0.76 | 0.62 | 0.47 | 0.83 |
|  | Livestock and Dairy Producers | 6121 | 1284 | 5108 | 5.7 | 0.44 | 0.53 | 0.49 | 0.36 | 0.67 |
|  | Mixed Crop and Animal Producers | 6130 | 777 | 3911 | 7.1 | 0.52 | 0.50 | 0.56 | 0.43 | 0.74 |
|  | Forestry and Related Workers | 6210 | 429 | 2638 | 6.8 | 0.67 | 0.79 | 0.59 | 0.39 | 0.91 |
|  | Deep-sea Fishery Workers | 6223 | 427 | 1658 | 6.1 | 0.50 | 0.54 | 0.47 | 0.32 | 0.70 |
|  | Biologists, Botanists, Zoologists and Related Professionals | 2131 | 1310 | 15289 | 9.2 | 0.89 | 0.95 | 0.78 | 0.65 | 0.94 |
|  | Environmental Engineers | 2143 | 1298 | 14019 | 7.8 | 0.75 | 0.81 | 0.75 | 0.61 | 0.92 |
|  | Graphic and Multimedia Designers | 2166 | 978 | 9635 | 8.9 | 0.79 | 0.76 | 0.77 | 0.62 | 0.95 |
|  | Refuse Sorters | 9612 | 518 | 3902 | 4.4 | 0.48 | 0.57 | 0.42 | 0.26 | 0.67 |
|  | Sweepers and Related Labourers | 9613 | 1381 | 13762 | 8.8 | 0.90 | 0.96 | 0.73 | 0.60 | 0.89 |
|  | Archivists and Curators | 2621 | 339 | 3528 | 4.4 | 0.45 | 0.43 | 0.46 | 0.28 | 0.77 |

^1^ Reference group: Employees with low likelihood of occupational SARS-CoV-2 exposure according to a Covid-19 job exposure matrix (sumscore for all eight rated indicators of SARS-CoV-2 workplace viral transmission = 0) ^6^

^2^ Adjusted for age (10 year groups)

^3^ In addition to age, adjusted for average number of RT-PCR tests and antigen tests in 4-digit-DISCO-08 codes (5 groups each)

^4^ In addition to age and test frequency, adjusted for duration of education at baseline (5 groups), number of hospital admissions for one or more of 11 chronic diseases in the 10 years preceding start of the pandemic (3 groups), country of birth (4 groups), geographical region (5 groups), number of household members (4 groups), probability of tobacco smoking (3 groups), bodymass index (2 groups), completed Covid19 vaccination (time varying variable, yes/no) and epidemic period (5 groups).
